# The interplay between neoantigens and immune cells in sarcomas treated with checkpoint inhibition

**DOI:** 10.1101/2023.05.20.23290277

**Authors:** Irantzu Anzar, Brandon Malone, Pubudu Samarakoon, Ioannis Vardaxis, Boris Simovski, Hugues Fontenelle, Leonardo A Meza-Zepeda, Richard Stratford, Emily Z Keung, Melissa Burgess, Hussein A. Tawbi, Ola Myklebost, Trevor Clancy

**Affiliations:** NEC OncoImmunity AS, Oslo Cancer Cluster, Ullernchausseen 64/66, 0379 Oslo, Norway; Institute for Cancer Research, Oslo University Hospital, N-0424 Oslo, Norway; Genomics Core Facility, Department of Core Facilities, Oslo University Hospital, Oslo, Norway; Department of Surgical Oncology, The University of Texas MD Anderson Cancer Center, Houston, Texas; Department of Medical Oncology, University of Pittsburgh Medical Center, Pittsburgh, PA, USA; Department of Melanoma Medical Oncology, The University of Texas MD Anderson Cancer Center, Houston, Texas; Department of Clinical Science, University of Bergen, Bergen, Norway

## Abstract

Sarcomas are comprised of diverse bone and connective tissue tumors with few effective therapeutic options for locally advanced unresectable and/or metastatic disease. Recent advances in immunotherapy, in particular immune checkpoint inhibition (ICI), have shown promising outcomes in several cancer indications. Unfortunately, ICI therapy has provided only modest clinical responses and seems moderately effective in a subset of the diverse subtypes. To explore the immune parameters governing ICI therapy resistance or immune escape, we performed whole exome sequencing (WES) on tumors and their matched normal blood, in addition to RNA-seq from tumors of 31 sarcoma patients treated with pembrolizumab. We used advanced computational methods to investigate key immune properties, such as neoantigens and immune cell composition in the tumor microenvironment (TME). A multifactorial analysis suggested that expression of high quality neoantigens in the context of specific immune cells in the TME are key prognostic markers of progression-free survival (PFS). The presence of several types of immune cells, including T cells, B cells and macrophages, in the TME were associated with improved PFS. Importantly, we also found the presence of both CD8+ T cells and neoantigens together was associated with improved survival compared to the presence of CD8+ T cells or neoantigens alone. Interestingly, this trend was not identified with the combined presence of CD8+ T cells and TMB; suggesting that a combined CD8+ T cell and neoantigen effect on PFS was important. The outcome of this study may inform future trials that may lead to improved outcomes for sarcoma patients treated with ICI.

## Introduction

Sarcomas are a rare heterogenous group of malignant bone and soft tissue tumors^1^ that account for 1% of all cancers but comprise 10-15% of solid tumors in children and young adults^2, 3^. Sarcomas are broadly divided into soft tissue sarcomas (STS) and bone sarcomas. The majority of sarcomas are STS, while malignant bone sarcomas account for 15% of sarcoma cases^4^. Bone sarcomas are most frequent in the younger population, with a median age at diagnosis of 15 years^5, 6^. Most sarcomas have a common mesenchymal origin and comprise a plethora of distinctive subtypes with more than 150 distinct categories recognized by the World Health Organization (WHO)^1, 7^. Each subtype is associated with diverse genetic, molecular and clinical features, making their comprehensive study, diagnosis and treatment challenging^8^. Moreover, many of these subtypes can occur at any age and are not restricted to any specific anatomical location. This heterogeneity and the rarity of sarcomas make the identification of mechanistic sensitivities and specific targeted therapies very challenging. However, the success of immunotherapy in diverse cancers and in stimulating anti-tumor immune activity may hold promise for patients with sarcomas.

Sarcomas have traditionally been classified as immunologically “cold” tumors, making them potentially challenging candidates for immunotherapy^9^. From a genome variation perspective, sarcomas can be classified in two major categories: with either simple or complex karyotypes^10–13^. In the first category, sarcomas harbor recurrent oncogenic translocations that may often drive tumorigenesis, accompanied by fewer other somatic genomic mutations or structural aberrations. This category is associated with a low tumor mutational burden (TMB). The second category has a higher but still moderate TMB and is distinguished by an imbalanced karyotype and increased genomic complexity, with high copy number variation and multiple structural aberrations^10, 14–16^. Because of the genomic instability associated with the sarcoma subtypes with complex genomes in the second category, they are more likely to exhibit higher levels of immune cell infiltration and so may be better candidates for immune checkpoint inhibition (ICI) threapy^17^. A recent study reported an improved immunogenicity due to the genetic instability in osteosarcomas^18^. However, this instability may also hinder a detailed understanding of the molecular mechanisms underlying carcinogenesis within the complex karyotype subtype. Osteosarcoma (OS), leiomyosarcoma (LMS), dedifferentiated liposarcoma (DDLPS) and undifferentiated pleomorphic sarcomas (UPS) are examples of this group^10, 11, 15^. Arguably, these relatively coarse subtype labels may not capture the genetic heterogeneity of individual tumors within each subtype. Nor do these subtype labels reflect the heterogeneity of subclasses within each subtype and their underlying tumor biology. Moreover, some simple karyotype sarcomas subtypes, such as chondrosarcoma, might evolve towards the second more complex category via the accumulation of secondary genomic alterations^19^.

Despite the extensive heterogeneity among sarcomas, patients with late-stage metastatic sarcoma commonly have a poor prognosis. Conventional cytotoxic systemic therapies are the standard-of-care, however effective treatment of metastatic sarcoma remains a challenge with five-year survival rates of barely 16%^20^. Cancer immunotherapy, particularly ICI therapy, has offered some major breakthroughs in recent years,^21^ and has revolutionized the treatment of some cancers including melanoma and renal cell carcinoma. ICIs have been the main drivers of cancer immunotherapy’s success in certain cancer indications. However, ICI monotherapy has durable clinical responses in only a minority of patients in certain cancer indications and is clinically ineffective in others. The only ICI therapy for sarcomas is the anti-PD-L1 atezolizumab, which gained Food and Drug Administration (FDA) clearance as of December 2022 for patients with unresectable or metastatic alveolar soft part sarcoma (ASPS)^22^. The use of ICI in advanced stage sarcomas has been reported in several clinical trials (including the SARC028/NCT02301039 that pertains to this study, where encouraging responses for a small number of patients have been previously reported^23–26^). In addition, several case reports have shown either a positive response or complete remission in patients treated with ICI^27, 28^. Several factors, including the wide variety of sarcoma subtypes and the varying expression of the checkpoint receptor in individual patients, may have impeded the identification of predictive biomarkers for ICI treatment response. The clinical trials have consistently reported an overall response rate (ORR) of roughly 15% with a median progression-free survival (PFS) of 3-4 months approximately, considering all histotypes^29–31^. DDLPS and UPS have exhibited slightly better responses while LMS and synovial sarcoma (SS) have persistently proven to be resistant to ICI monotherapy^29, 30^. An immunologically “cold” TME may be a limiting factor in achieving significant ICI therapy clinical success from ICI therapy in most sarcomas. The potential benefits of modulating the TME with neoadjuvant chemotherapy towards a “hotter” or more active immune status and hence, becoming more susceptible to ICI, have been reported for OS^32^. A significant improvement in the clinical benefit rates of ICI was observed when stratifying specific patients according to TME, tertiary lymphoid structures (TLS)^33–35^, histological type^36, 37^, and PD-L1 expression^30, 38, 39^. Additionally, the role of TMB in sarcoma, which has previously been shown to be relevant in many cancer types as a predictive biomarker to ICI^40^, is still unclear and is being investigated further^41^. Since most sarcomas have relatively low TMB^42^, they do not commonly present a sufficiently high frequency of non-synonymous mutations to produce enough tumor-specific antigens (neoantigens). A lack of targetable neoantigens may reduce the chance of an effective antitumor immune response after ICI treatment. The high frequency of sarcoma fusion events, on the other hand, provides an appealing source for targetable neoantigens to unlock immunotherapeutic response^43^, although the expression of the gene fusion products needs to be evaluated^44^.

The few positive clinical responses to ICI among sarcoma patients, however, motivate further research to decipher the mechanisms of immune escape or resistance to therapy, including the interplay between neoantigens, immune cells within the TME, and other factors that may guide sarcoma patient selection, and thereby optimize clinical outcome^42, 45^. In the SARC028 study, durable responses were observed in about 20% of STS, especially in patients with UPS and DDLPS, whereas bone sarcomas had a modest response rate. Since the initial trial had limited numbers of patients for each subtype, an expansion cohort of patients with UPS and DDLPS was included and which showed somewhat lower clinical response rates^46^. Another study investigated a histology-agnostic immune gene expression signature (Sarcoma Immune Class, SIC) on STS and showed that this was both predictive of response to ICI and prognostic of survival in the SARC028 patient cohort. Importantly, the SIC class that exhibited the greatest response to ICI (SIC E) and best prognosis was characterized by the presence of intratumoral TLS and B-cell gene expression signature^33^.

In this study, we investigated samples from patients in the SARC028 trial diagnosed with the following bone sarcomas: OS, chondrosarcoma (CS) and Ewing sarcoma (ES); and the following STS: UPS, DDLPS, LMS and SS. We performed whole-exome sequencing (WES) of tumor and matched normal blood from patients and RNA-sequencing (RNA-seq) of tumor to identify neoantigens corresponding to multiple sources of genomic variants including single nucleotide variants (SNVs), small insertion and deletions, and gene fusion events. The RNA-seq data from tumors was also used to characterize immune cell infiltration into the TME. We also explored the expression patterns of a collection of immune-related genes to improve our understanding of the immunobiology of ICI-treated sarcomas. The subsequent exploratory multivariate survival analyses revealed that the specific context of the immune cell composition of the TME and its interplay with immunogenic neoantigens may be important for improved PFS. The insights gained from this analysis will aid the identification of prognostic biomarkers underlying disease mechanisms and will be informative in future clinical trial designs and studies of ICI therapies in sarcomas.

## RESULTS

### Immune cell infiltration patterns in sarcoma patients treated with ICI therapy

We first analyzed tumor expression profiles by RNA-seq of bulk tumor samples (see Methods). A principal component analysis (PCA) suggested a clustering of patients that corresponded to the sarcoma subtypes (Supplementary Figure S1a). Certain subtypes, such as LMS, had a notable within-subtype heterogeneity, while other subtypes such as SS, were more similar. A subsequent PCA focused exclusively on the selected collection of immune-related genes (Supplementary Figure S1b) suggested that a distinctive difference in immunogenomic expression profile may exist between the different sarcoma subtypes treated with ICI (Supplementary Figure S1b). Interestingly, two out of the three patients responding to ICI clustered together in both PCA plots.

A trend towards elevated expression of genes related to the immune response in UPS and OS relative to the other sarcoma subtypes was observed when using hierarchical clustering of immune-related genes as illustrated in Figure 1a (with an average transcript per million (TPM) of 94 and 84, for UPS and OS respectively, compared to the other sarcoma subtypes where the average TPM was consistently less than 60). The samples made up two main clusters, A, with almost all OS, and B, with all SS, suggesting some relation to the karyotypic subclasses. Further, a hierarchical clustering of the predicted TME composition is depicted in Figure 1b, and in Figure 1c a bar chart of the specific immune cell fractions of each cluster from Figure 1b is portrayed. Cluster 3 in Figure 1c consisted mostly of OS and seemed to be dominated by an elevated level of macrophages (both M1 and M2 types were detected). Interestingly, Cluster 4 was enriched in monocytes, and most of the patients in that cluster had a stable disease (SD) response to ICI therapy. Cluster 5 seemed to be the “coldest” immunological group relative to the other clusters, not surprisingly harboring most of the ES samples driven by specific gene fusions. Interestingly, B cell infiltration into the TME was predicted to be a common sarcoma feature at relatively consistent levels across all sarcoma subtypes (Figure 1c).

**Figure 1.**
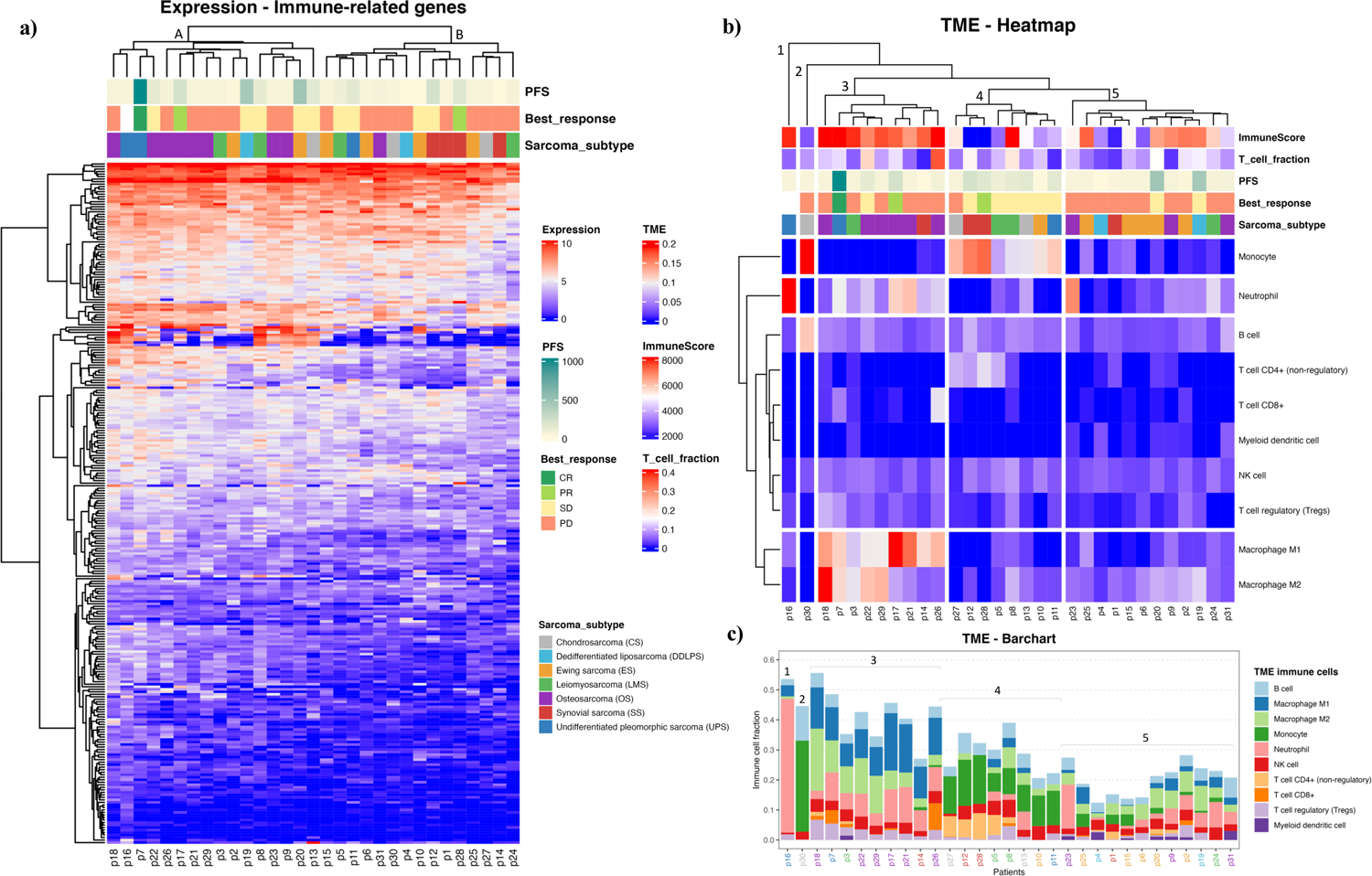
**a)** Heat map representing the hierarchical clustering analysis of all baseline samples in two main clusters considering the expression profiles of the immune-related genes. (Progression free survival (PFS) is capped at 1000 days). **b)** Tumor microenvironment (TME) clustering using the fraction of each immune cell type per sarcoma sample predicted by quanTIseq. ImmuneScore values are based on ESTIMATE analysis. **c)** TME bar chart with immune cell fractions. Patient IDs are colored by sarcoma subtype as in a) and b). The clusters from b) are indicated.

As observed in Figures 1b and c, CD8+ T cell infiltration was also predicted at varied levels among the sarcomas. The T-cell infiltration predicted by quanTIseq was validated by the TcellExTRECT tool. As can be observed in Figure 1b, TcellExTRECT results were concordant with quanTIseq, with a positive significant correlation of 0.41 and p-value = 0.022. The generic infiltration levels of non-cancer cells (*i.e.,* stroma and immune cells) across the different sarcoma samples were also evaluated using the ESTIMATE toolkit. The stromal scores from ESTIMATE ranged from 1977 to 7910, while the ESTIMATE immune cell infiltration scores ranged from 574 to 9821, and finally the ESTIMATE tumor purity scores ranged from 2959 to 11086. Regarding the sarcoma subtypes, the mean and standard deviation of immune scores sorted from highest to lowest were as follows: OS, 6870 (1626); DDLPS, 6295 (2652); UPS, 60797 (2920); LMS, 5977 (2268); ES, 5832 (1884); CS, 4139 (1201); and SS, 2358 (2838). These ESTIMATE predictions corroborated the observations in Figure 1, in that OS had a higher immune activation compared to the other sarcoma subtypes. In Figure 1b, the “ImmuneScore” annotation bar represents the results of the ESTIMATE tool, where it can be clearly observed that Cluster 3, consisting mostly of OS, was the cluster with highest immune scores.

Additionally, we performed a differential expression (DE) analysis comparing the patients with a clinical response to ICI (responders) to the other patients. We identified a total of 727 differentially expressed genes (DEGs), of which 209 were up- and 518 down-regulated, with an absolute fold change larger than 1 and a p-value less than 0.05 (Supplementary Figure S2a). Due to the small number of responders (three), multiple test correction was not applied. Enrichment analysis of the up-regulated genes revealed a significant enrichment of several immune-related Gene Ontology (GO) terms (Supplementary Figure S2b, corroborating the notion that an immunologically active or “hot” TME leads to improved clinical outcome to ICI therapy^33^). A detailed table with all DEGs and the complete list of up-regulated GO terms is provided in Supplementary Table 1 and 2, for DEGs and GO terms respectively. The study focuses on immunogenomic properties of ICI-treated sarcoma patients; hence cancer hallmarks were not examined.

### Sarcoma tumors exhibit a highly heterogeneous neoantigen landscape

Using a state-of-the-art somatic mutation calling framework^47^, we inferred a comprehensive mutational landscape of the 31 baseline sarcoma tumor samples (see Supplementary Figure S3 for a detailed overview). Each 9mer and 10mer peptide that had a somatic mutation was matched to the personalized HLA genotype of each individual patient to identify immunogenic neoantigens likely to be presented by the patient’s HLA alleles on their tumor cell surface. Such high-quality neoantigens were predicted using the NEC Immune Profiler (NIP) software (see Methods)^48^, that uses an integrated artificial intelligence (AI) approach trained on proprietary data to predict antigen presentation (AP) scores, which can range from 0 to 1, for each candidate neoantigen. The distribution of neoantigen load (NAL) (see Methods) with respect to each sarcoma histological subtype was then assessed (Figure 2a) and revealed a highly heterogeneous NAL both between and within subtypes, ranging from 0 to 206 for intra-subtypes and median range of 15.0 to 112.0 for inter-subtypes. The DDLPS, UPS and LMS subtypes exhibited the highest NAL overall; the striking score for DDLPS is due to one case, p4, who had an extremely high number of gene fusions (Figure 2b). 461 (28.9%) of the mutated peptides had an AP score (see Methods) greater than 0.5 and were also predicted to bind more than one HLA allele in the same patient. Figure 2b shows the number of neoantigens generated in each patient, stratified by the type of somatic mutation, outlining a broad diversity of potential neoantigens detected within each subtype.

**Figure 2.**
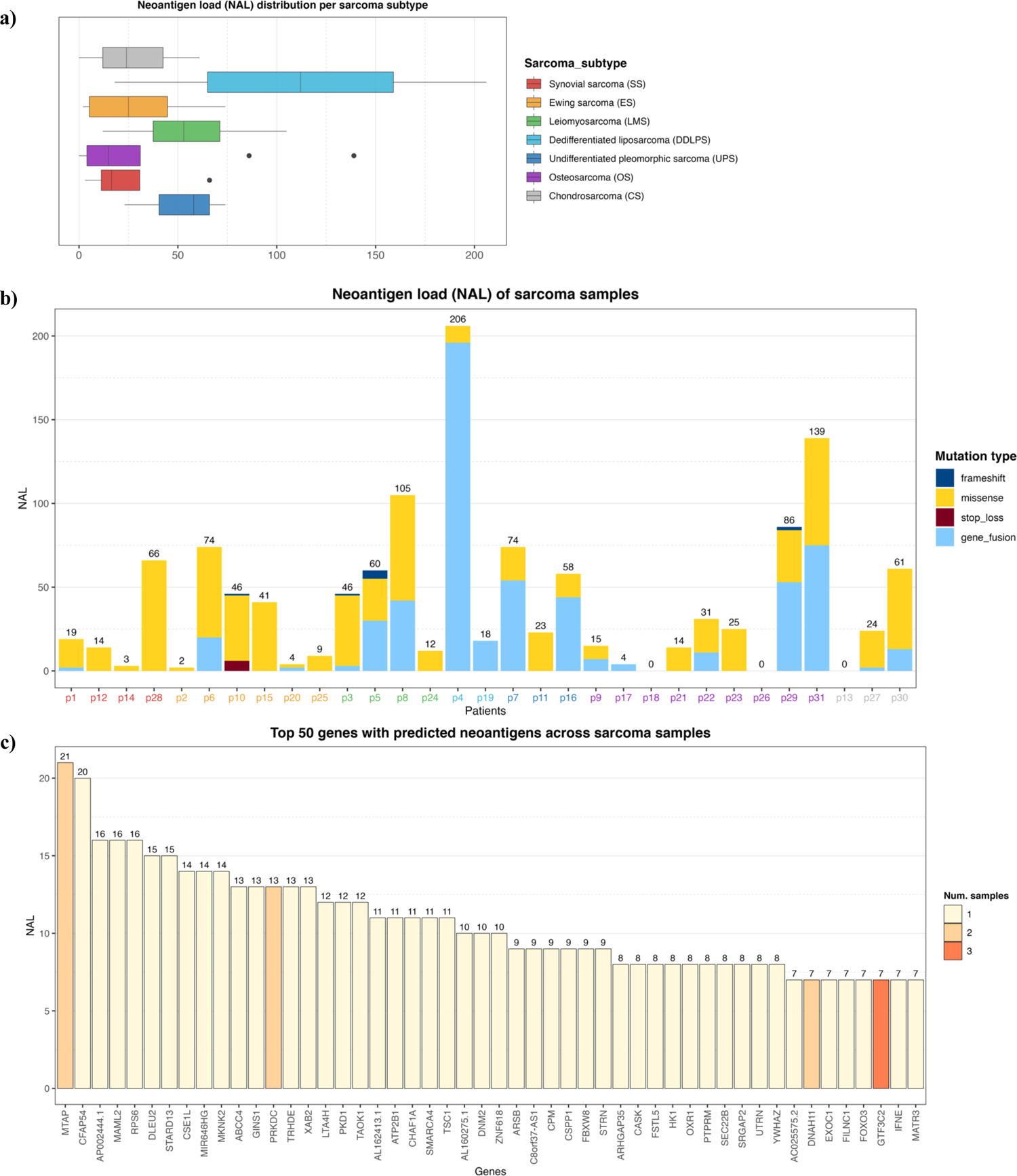
**a)** Neoantigen load (NAL) distribution across the different sarcoma histological subtypes. The lines inside each box represents the median NAL value for each subtype while the dots are outliers. **b)** NAL profiling of sarcoma samples and the contribution of each somatic variant type to NAL. Patient IDs are colored by sarcoma subtype as in a). **c)** Top 50 genes with predicted neoantigens across sarcoma patients. The colors represent the number of patients with neoantigen candidates arisen from the given gene.

In this study, no correlation between a conventional TMB and NAL was found (Spearman Rank correlation coefficient of −0.08, and p-value of 0.66). The conventional TMB calculation typically does not take gene fusions into account, and as can be observed in Figure 2b, gene fusions were one of the dominant contributors to NAL for several patients. However, a clear positive correlation between the number of gene fusions and NAL was observed, with a correlation coefficient of 0.92 and p-value of 2.65e-13. We also measured the contribution of each gene to the overall NAL across all the patients. Figure 2c shows the top 50 most frequently mutated genes that gave rise to candidate neoantigens. For instance, the gene MTAP, a methylthioadenosine phosphorylase, known to be deficient in some tumors^49^, contributed 21 neoantigens (Figure 2c) across two patients (Supplementary Figure S4). The PRKDC gene, encoding for a DNA-dependent protein kinase catalytic subunit, is involved in DNA repair, the establishment of immune tolerance, and genome stability, and thus, has the potential predictive biomarker for ICI therapy^50^. We detected 13 neoantigens across two patients for the PRKDC gene (Figure 2c and Supplementary Figure S4). No shared neoantigens were found between the different patients, in which is similar to other studies^51^ and expected here due to the highly heterogenous mutational landscape across sarcoma subtypes.

We next analyzed the distribution of the ranked AP scores of the top ten neoantigen candidates for each of the baseline samples. The patients were pooled according to their clinical response, consisting of three responders, 18 patients with progressive disease (PD), and 9 patients with stable disease (SD). The distribution of AP scores, and the means of the populations between each group, was then compared using the Welch’s test (Figure 3a). A marginally significant difference in neoantigen scores between the responders and PD groups was observed (p-value = 0.078). There was no significant difference between the responders and SD groups (p-value = 0.63), but the comparison between the SD vs PD groups emerged with a significant difference (p-value = 0.001). Additionally, also using the Welch’s test, we compared the PD group against the remaining non-PD groups (*i.e.,* SD and responders pooled together), whereby a significant difference emerged from the analysis (p-value = 0.001) (Figure 3b). Interestingly, this analysis revealed, on average, a higher neoantigen quality for patients without PD.

**Figure 3.**
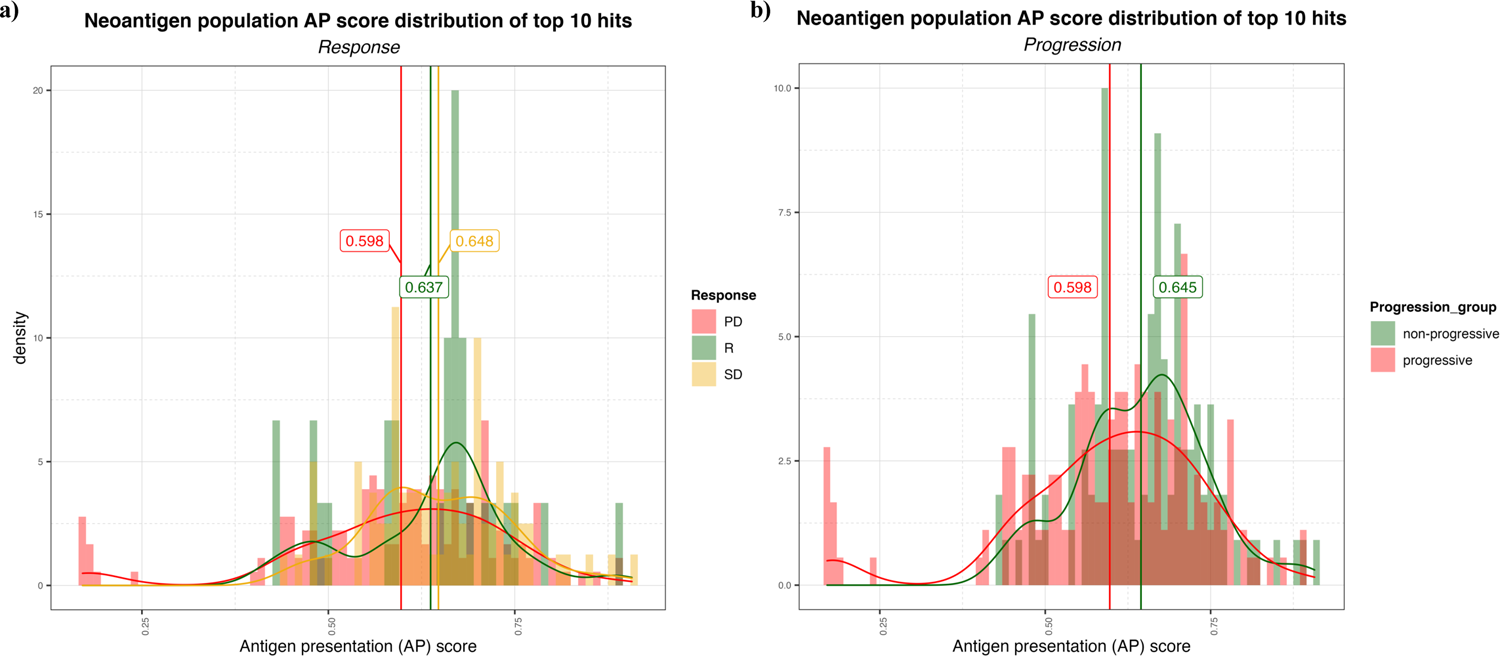
**a)** Distribution of the AP scores for the top 10 (ranked by AP score) neoantigen candidates for progressive disease (PD), stable disease (SD) and responder (R) group. The vertical lines represent the mean AP score for each group. **b)** Distribution of the AP scores for the top ten (ranked by AP score) neoantigen candidates for non-progressive (SD + R) and progressive (PD) groups.

For this specific study, there were unfortunately too few patient samples available for analyses post ICI therapy to derive significant conclusions. For the few samples that were available post therapy; none responded to ICI. In addition, removal by immunoediting of somatic mutations or immunogenic neoantigens caused by ICI therapy was not detectable among the few available ICI-treated samples (Supplementary Figure S5).

### Survival and clinical response of TMB, NAL and immune cell TME in ICI treated sarcomas

We next evaluated the effect of various immunogenomic features on PFS of the patients using Kaplan-Meier (KM) survival analysis. Conventional thresholds such as mean/median or Maximally Selected Rank Statistics failed to yield significant results in most cases probably due to the considerable diversity of the dataset consisting of seven sarcoma subtypes and only 31 samples. Hence, we used an exploratory optimal binning approach (see Methods) to group patients into two (low and high) groups for univariate analyses (*i.e.,* each immunogenomic feature was analyzed individually); for bivariate analysis, the univariate thresholds were used to stratify patients into four groups (*i.e.,* the immunogenomic features were analyzed in pairs of low/low, low/high, high/low, high/high). We note that the small sample sizes available in this study may limit the robustness of the log-rank test associated with the KM analysis. We also did not account for multiple hypothesis testing in this analysis. Consequently, we do not make strong claims of statistical significance in this preliminary study. Nevertheless, this exploratory analysis does reveal features associated with differences in PFS among patient groups and which could guide further avenues for future studies.

We first performed a statistical interrogation of the immune cell infiltration in to the TME in a univariate analysis. We identified a signal that an elevated fraction of infiltrated T cells into the TME as measured by TcellExTRECT tool (see Methods), led to improved PFS with a log-rank p-value in the KM analysis of 0.00096 (Figure 4a). The group of patients presenting a higher proportion of infiltrated T cells had a median PFS of 173 days while the lower group had a median of 48 days. Figure 4b demonstrates the suitability of the supervised optimal binning approach to find the best threshold to generate the different groups evaluated in the KM survival analysis. A window of low p-values across different thresholds indicates a more robust separation of low and high groups which is not sensitive to the exact chosen threshold. Improved PFS was also observed to be associated with increased levels of macrophage M1 and M2 cell infiltration (log-rank p-values 0.047 and 0.019 for M1 and M2 cells respectively, Supplementary Figure S6a and S6b) and associated with B cells with a log-rank p-value of 0.074.

**Figure 4.**
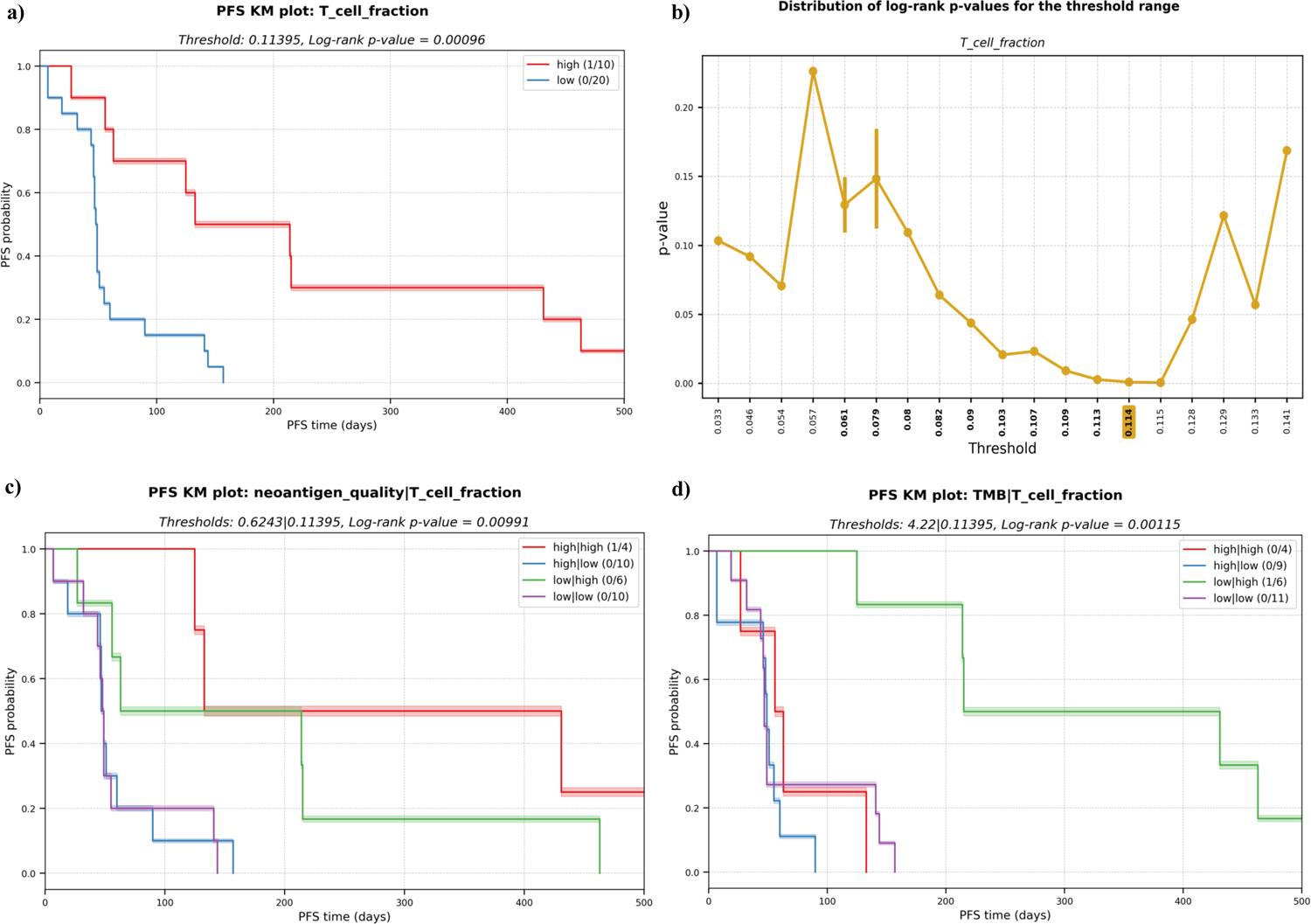
**a)** Kaplan-Meier (KM) plot for T cell fraction univariate analysis. A higher fraction of tumor infiltrated T cell leads to better progression-free survival (PFS). PFS capped to 500 days for visualization purpose. **b)** Distribution of log-rank p-values for the different thresholds evaluated during the exploratory supervised optimal binning approach. In bold, all the thresholds resulting in groups of at least ten patients each, among those, the one with lowest p-value (highlighted in yellow) is selected and used in a). **c)** KM plot for the multivariate analysis with the combinatorial effect in PFS of neoantigen quality and T cell fraction; and **d)** TMB and T cell fraction.

To determine the predictive potential of the effect of combining certain immunogenomic properties for prediction of PFS, we performed multivariate analyses. We observed that a higher neoantigen quality (see Methods) combined with a high infiltrated T cell fraction was associated with an improved PFS (median of 282 days) with a log rank p-value of 0.006 (Figure 4c). High T-cell infiltration with low neoantigen quality was still associated with good PFS (median PFS = 139 3 days;). However, this PFS is lower than the median PFS among all patients with a high T-cell fraction (173 days); this highlights that incorporating neoantigen quality with T cell fraction improves prognostic patient groupings compared to using T-cell infiltration alone. Low T-cell fraction was associated with poor PFS, regardless of whether there was high neoantigen quality (median PFS=49 days) or low (median PFS=48 days; Figure 4c). The same association with improved PFS was not observed with TMB and T cell fraction. In fact, high TMB and high T cell fraction was associated with poor PFS (median PFS = 59 days) while patients with low TMB and high T cell fraction had favorable PFS (median PFS = 215 days) (Figure 4d). A similar pattern was observed when combining TMB and ESTIMATE score (log-rank p-value 0.01) with the longest PFS reported for the group with low TMB and high immune score (Supplementary Figure S7a). This contrasted with the neoantigen quality, where a minimal or low level indicated a longer PFS time if in the presence of high T cell fraction (see Figure 4c). Furthermore, the presence of several immune cells was also found associated with better PFS, where the absence of both (low/low) was often correlated with the worst outcome (Supplementary Figure S7).

### Investigation of immune escape parameters: antigen presentation machinery, personalized HLA-typing, and the tumor-specific HLA status

The polymorphic nature of HLA alleles and its association to tumor-immune escape called for accurately typing and evaluating the mutation and expression status of each HLA allele in the patients^52^. The status of the HLA locus in the different sarcoma subtypes was evaluated using NeoOncoHLA (see Methods). Using this personalized HLA-typing approach, the somatic mutations and tumor specific expression of each patient-specific HLA allele was described. A total of 18 somatic variants affecting HLA class I alleles were detected among 14 patients (45% of the patients presented at least one somatic mutation affecting one HLA allele). Seven of those 18 were non-synonymous variants, and six of those seven affected the peptide binding regions of the alleles. Figure 5 depicts the expression of each HLA allele estimated as TPM and the NAL associated with each sample’s HLA-A and HLA-B alleles. The number of good neoantigen candidates for the HLA-A*02:01 allele in p9 increased from nine at baseline to 24 in week-8, while its expression (measured as TPM) decreased from 861 to 350. This could indicate a possible immune escape mechanism in this patient, through the downregulation of this HLA allele’s expression (see Figure 5). Although p31 showed numerous good neoantigen candidates, the expression of the HLA alleles seemed to be slightly downregulated, particularly for HLA-A*11:01. We did not observe this putative immune escape pattern consistently for HLA status as a global trend across all patients, and we had too few treated samples to draw conclusions. HLA expression did not demonstrate statistical significance in the survival study. Nonetheless, in the multivariate KM survival analysis, we found a significant separation for HLA-C (log-rank p-value = 0.001) expression in conjunction with the predicted NK cell fraction. Patients with a high NK cell infiltration combined with low HLA-C expression had the best PFS (median PFS = 157 days), while a high NK and HLA-C values resulted in short survival (median PFS = 50 days) (Figure 6a). This trend was not observed when combining HLA expression with T cell infiltration into the TME, where the high HLA expression and high T cell infiltration group resulted in similar PFS times as low HLA expression and high T cell infiltration group (see Figure 6b, c and d).This pattern could theoretically be owed to the combined benefit of enhanced NK cell activity due to the decreased HLA-C expression and the increased presence of T cells in the TME modulating tumor cell killing through HLA-A and/or -B antigen presentation to T cells; in the backdrop of improved PFS with high neoantigen quality with high T cell fraction reported above (see Figure 4c).

**Figure 5.**
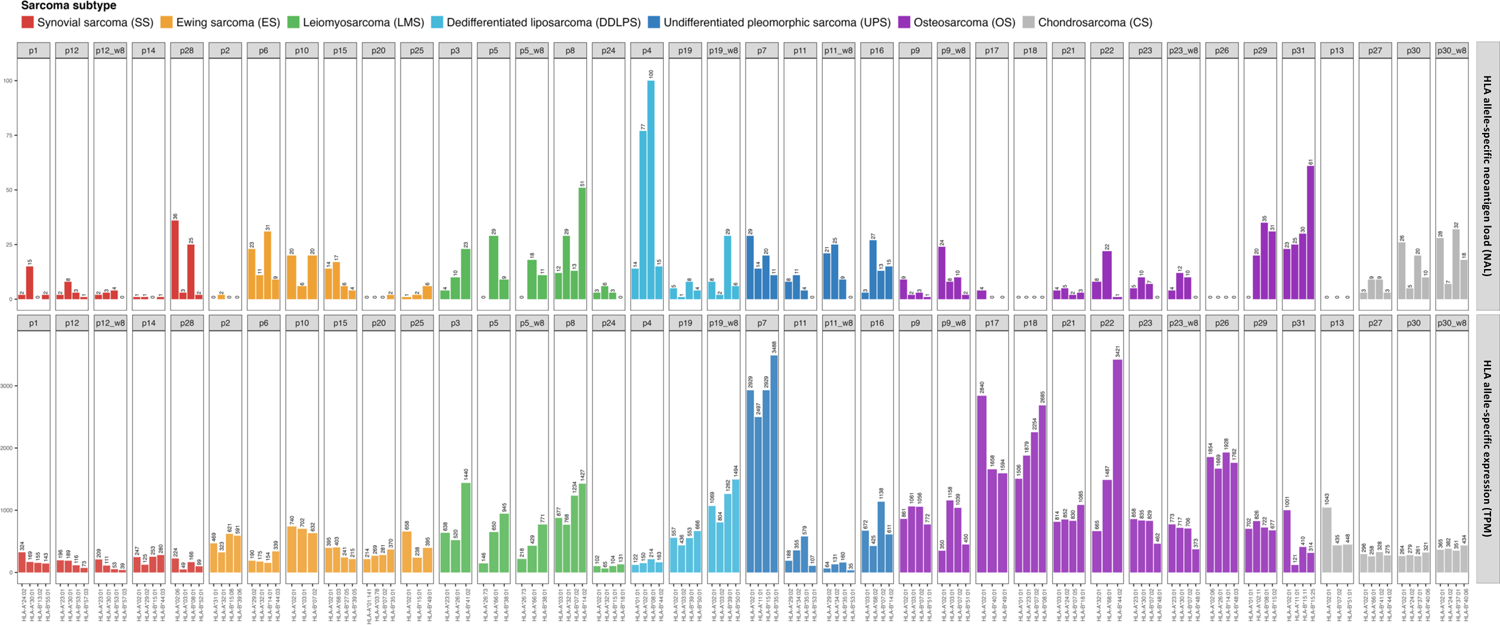
HLA-A and -B allele-specific expression calculated as TPM across the different ICI-treated sarcoma samples (bottom histogram) and the correspondent NAL for each allele (top histogram).

**Figure 6.**
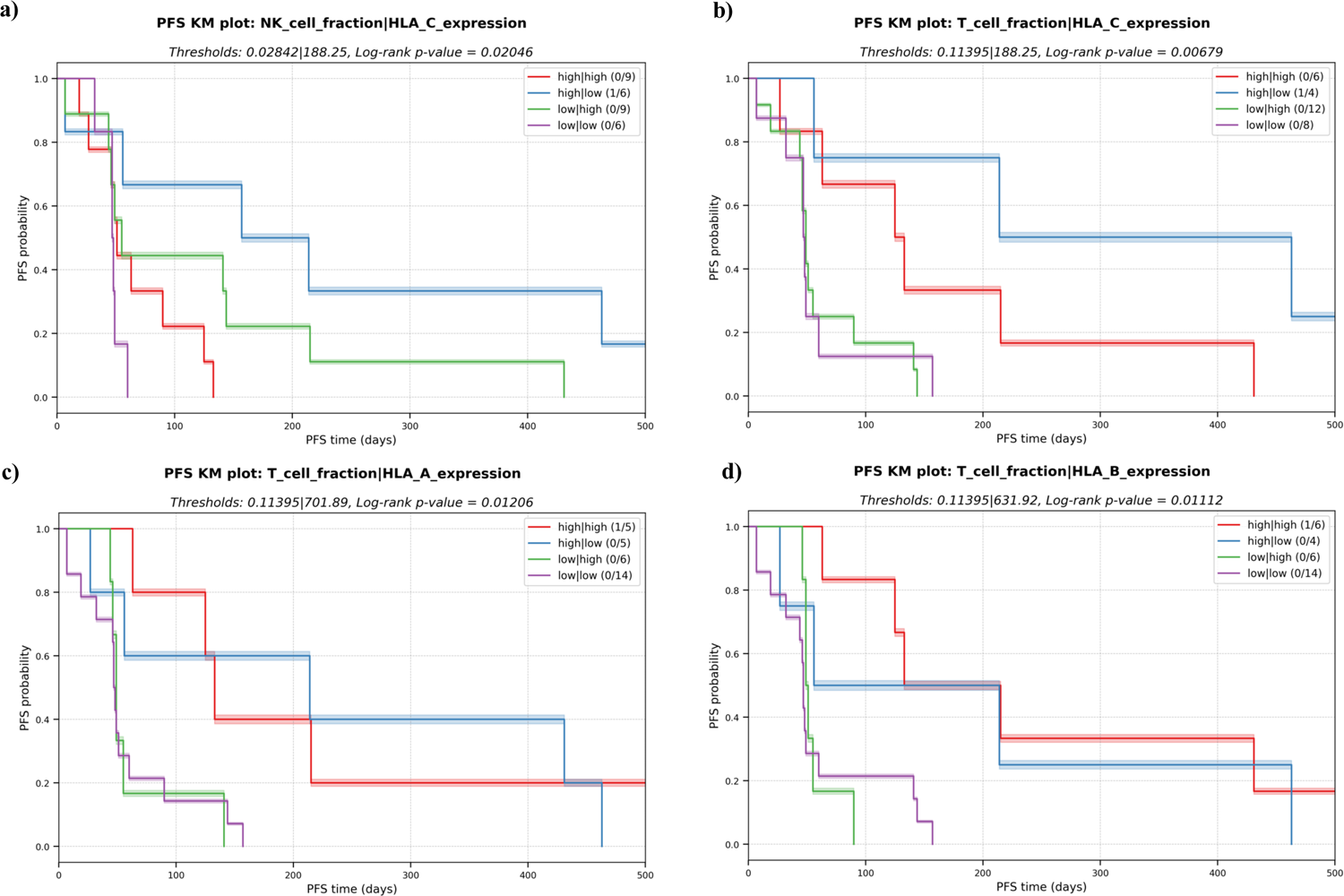
Kaplan-Meier (KM) plots for the multivariate analysis with the combinatorial effect in PFS of HLA expression and immune cell fractions in the TME. **a)** NK cell fraction and expression of HLA-C. **b)** T cell fraction and expression of HLA-C. **c)** T cell fraction and expression of HLA-A. **d)** T cell fraction and expression of HLA-B.

Importantly, the expression profile of genes linked with the antigen presentation machinery (APM), in addition to HLA expression, revealed probable patterns of immune evasion in sarcoma. Among the APM genes we profiled (see Methods), we found that a decreased expression of beta-2-microglobulin (B2M), MHC class II transactivator CIITA endoplasmic reticulum aminopeptidase 2 (ERAP2), transporter 2 (TAP2) TAP binding protein like (TAPBPL) and MHC class II transactivator were significantly associated with a shorter PFS (Figure 7). Consistent with previous research, these findings underline the utility of APM profiling as potential biomarkers of tumor cell antigen presentation status and immune escape^53–55^.

**Figure 7.**
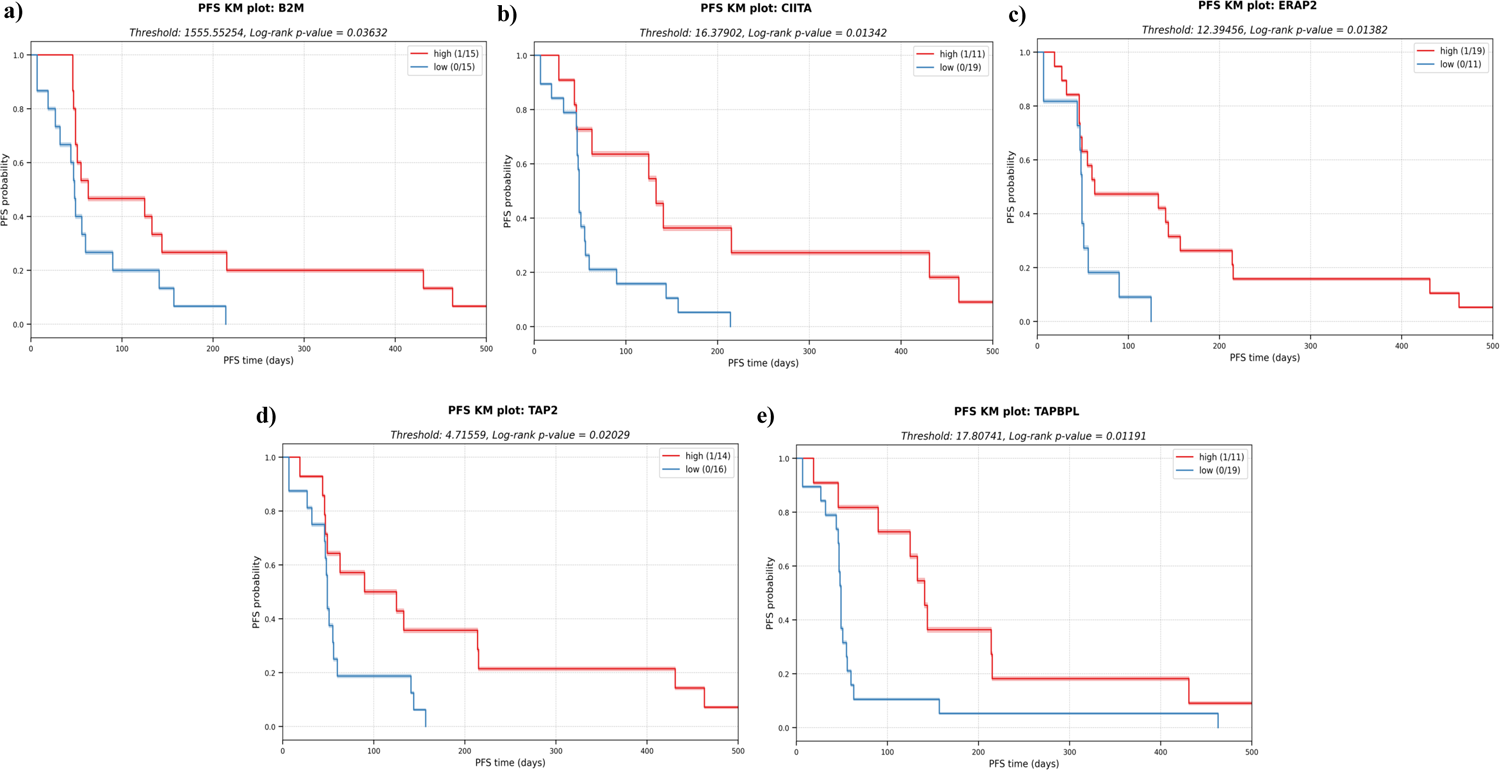
KM plots for several APM components analyzed in a univariate analysis. The downregulation of these components in the tumor cells might impair antigen presentation and subsequent escape from immune system. **a)** Beta-2-microglobulin (B2M). **b)** MHC class II transactivator CIITA. **c)** Endoplasmic reticulum aminopeptidase 2 (ERAP2) **d)** Transporter 2 (TAP2) **e)** TAP binding protein like (TAPBPL).

## Discussion

The efficacy of pembrolizumab in patients with advanced bone and soft-tissue sarcoma (STS) was assessed in the SARC028 trial (NCT02301039) and demonstrated that only some of the many different histological subclasses of sarcomas had a positive clinical response to ICI^56^. In general, the results were modest, with promising results within some of the STS subtypes^23^. Many studies have reported the importance of immune cell infiltration into the TME and their complex interplay with tumor cells in the clinical outcome of cancer patients^12, 29, 57^. Furthermore, in recent years, neoantigens have emerged as important immunotherapy targets that play a central role in the HLA-restricted T-cell response and have been linked to the clinical efficiency of ICI therapy in several cancer indications^58, 59^. Moderate responses have been found in clinical trials of ICI in sarcomas, and only a subset of patients has benefited with durable clinical outcomes^56^. Due to this modest response to ICI therapy, comprehensive immunogenomic analysis of sarcomas guided by state-of-the-art AI tools to predict neoantigens and their interplay with respect to immune cells in the TME was warranted in this study to help elucidate some of the mechanism of possible immune escape and resistance of sarcomas to ICI therapy and to enable prior identification of patients most likely to respond.

We used a series of computational and AI methods to investigate several tumor immunogenomic properties, including the somatic mutational landscape, neoantigens, expression of key immune-related genes (e.g., APM genes), and the TME immune cell composition from the available SARC028 trial samples. The analysis was conducted on NGS data from 31 sarcoma patients, comprising WES from matched tumor and blood (germline) samples and tumor RNA-seq data. In terms of ICI response, three patients had a clinical benefit from pembrolizumab (one complete responder and two partial responders), nine had stable disease (SD), 19 had progressive disease (PD), and one did not have response information available.

Using the RNA-seq data from the tumor samples, a DEG analysis revealed several immune-related GO terms were significantly enriched in the responders, suggesting that an immunologically active TME may lead to an improved clinical response to ICI therapy. A further interrogation revealed three distinct TME immune profiles based on hierarchical clustering of the tumor transcriptome data; (1) an immune active cluster (cluster 3), formed mostly by OS, and enriched in macrophages and CD8+ expression, (2) a cluster enriched by monocytes and with patients with SD response (cluster 4), and (3) a slightly immune desert cluster consisting of all ES patients except one. Importantly, two of the three ICI responders, including the patient experiencing a complete clinical response, were included in the TME immune active group (cluster 3). This was consistent with the differential expression analysis which showed that responders are associated with a pre-existing immune activity in the TME. The RNA-seq data was also used to deconvolute the immune cell composition in the TME, and then applied to a univariate KM survival analysis to identify immune cell infiltration patterns associated with improved PFS. We observed that higher fractions of T cells and macrophages were all associated with longer PFS. These observations were consistent with previous studies that examined the importance of the immune cell TME for the ICI response in sarcomas^60^, although contradictory results have been reported in the literature. For example, B-cell signatures correlating with longer survival times^33^, whereas the converse has been reported for CD8+ T cells^39, 61–63^. Additionally, our observation of elevated macrophages in OS was consistent with previous studies^32^. Similarly, using the RNA-seq data, we found that a lower expression of APM-related genes, including B2M, CIITA, ERAP2, TAP2 and TAPBPL, was also associated with a shorter PFS, reflecting the increasing interest in APM biomarkers for clinical outcome and immune escape in cancer^64^. Finally, the RNA-seq data revealed a pattern whereby a decreased allele-specific HLA-C expression, combined with high NK cell infiltration, was linked with improved PFS. This is consistent with the well-established trend of downregulation of class I HLA allele expression correlating with NK cell activity^65^.

The somatic mutation profile of the sarcomas was highly variable both within and between histological sarcoma subtypes. OS samples had the highest TMB score, followed closely by UPS, with CS having the lowest and least variable TMB. While many genes harbored mutations across the different patients, the top 4 most commonly mutated genes were all members of the mucin (MUC) gene family, known to play a role for epithelial tissues and reported in some neoplastic lesions^66^. However, this trend may be explainable by the high degree of exon repeats in the MUC family of genes among individuals^67^. In addition to small variants, copy-number variations and chromosomal rearrangements were found in many patients, including known ES driver gene fusions of EWSR1 and SS18 genes. An interesting observation here was the high number of fusion-generated neoantigens in DDLPS compared to the other subtypes. The complex karyotypes of DDLPS would be expected to contain many genome rearrangements, but so would OS and UPS, which do not show this pattern. This observed difference is challenging to explain but could contribute to the better response seen in patients with DDLPS compared to OS but does not explain the better response of UPS cases^30^.

Increased neoantigen load (NAL) has previously been positively correlated with TMB^68, 69^, and therefore described to correlate with ICI therapy response^70–72^ in certain cancer indications. In this study, however, we did not find a correlation between TMB and NAL. This finding reinforces the argument that higher TMB does not necessarily always equate to higher NAL. It is important to note that the conventional TMB calculation does not consider gene fusions and therefore bypasses powerful sources of neoantigens. Additionally, the conventional TMB calculation does not consider the key determinants of antigen presentation or immunogenicity, such as antigen processing, HLA binding and the expression of the somatically mutated peptides, in addition to their distance from self (that is, the “wild type” protein). we revealed a potential “bad, good, bad” pattern for PFS for “low, medium, high” TMB, respectively. One can postulate that the low TMB may reflect a relatively cold tumor harboring few immunogenic neoantigens; other work has also shown that ICI is often not effective for such cold tumors, so we may expect a low PFS for such patients ^73^. On the other extreme, a high TMB may indicate a very complex tumor which already manifests a variety of immune escape mechanisms or harboring many neoantigens that are subject to a high degree of peripheral T cell tolerance, and is therefore unlikely to respond to immunotherapy ^74^. However, we postulate here, that there is an argument supporting the notion that medium TMB may lie in a potential neoantigen “Goldilocks zone” in which the tumor expresses and presents neoantigens harboring sufficient differences from self that an ICI stimulated immune system can recognize and respond to the tumor, while simultaneously the mutated tumor genome is not yet so complex as to allow for immune escape.

In our univariate analysis, neither NAL nor neoantigen quality were associated with improved PFS. However, when combining neoantigen quality with the presence of T cells, we observed a striking joint behavior associated with longer PFS compared to the presence of T cells alone. When the T cell fraction was low, PFS was always low (median PFS=48 days) regardless of the neoantigen quality. This was consistent with the finding that immune T cell deserts are not associated with good outcomes for ICI therapy^75^. Additionally, it is reasonable that high-quality neoantigens are not effective if there are no T cells in the surrounding environment to recognize them. In the case of a high T cell count but low quality neoantigens, PFS was modestly improved (median PFS=139 days) compared to the low T cell count patients. Remarkably, patients with both a high CD8+ T cell count and high-quality neoantigens had improved PFS (median PFS=282 days). The observed improved PFS when combining the tumor infiltrated CD8+ T cell fraction and neoantigen quality was not replicated when combining T cell fraction and TMB. In this case, a good PFS was observed only for low TMB with high T cell fraction, suggesting that the quality and not quantity of neoantigens might be more relevant in certain settings for clinical benefit^76^. Overall, the comparison of neoantigen load vs TMB in the context of T cell infiltration indicated that the AI neoantigen prediction platform used to identify neoantigens is reliably predicting mutated peptides presented on the tumor cell surface that are potentially immunogenic. For B cells, a similar picture emerged, except it was required for both the neoantigen AP scores and the B cell fraction in the TME to be elevated for longer PFS. This finding is reflective of the new landscape emerging on the importance of B cell tumor infiltration, prognosis, and response to immunotherapy^33^. Overall, the KM survival threshold analysis findings in this study were also consistent with the TME clustering which found that responders were associated with a pre-treatment, immune-inflamed TME.

The KM survival analysis based on the supervised optimal binning approach we used to arrive at the insights described in this study has several limitations and caveats. First, each set of thresholds could be considered as a hypothesis in the sense of statistical testing; appropriate multiple test corrections, such as Benjamini-Hochberg would ideally be applied if the p-value was to be interpreted for true statistical significance. In this exploratory work on a limited number of samples, though, we simply aimed to identify the best bins for the data and did not intend to make broad robust statistical claims. Thus, we believe the approach is still justified as it provides informative insights from a small patient cohort, but we acknowledge the need to validate the results in larger patient cohort. Secondly, some choices of thresholds would have resulted in very small groups; the assumptions of many statistical tests do not hold in such cases. In this work, we limited this problem by only considering univariate thresholds which result in at least ten individuals in all groups.

To the best of our knowledge, this is first study to exhaustively profile the immune cell TME of sarcomas with its interplay with immunogenic neoantigens under the context of ICI therapy, in a manner that uses advanced computational AI tools to comprehensively capture this important interplay. While the sample size for this study was small, the insights gained were suggestive that the interplay between neoantigens and immune cell infiltration patterns into the TME is a key prognostic marker of clinical response to ICI and PFS. This therefore warrants further clinical and biomarker studies with larger sarcoma cohorts.

## Material and methods

### SARC028 trial cohort data description

The SARC028 trial (NCT02301039) recruited 86 patients. Of those, 80 patients (40 with advanced STS and 40 with bone sarcomas) were eligible for the ICI therapy (pembrolizumab, an anti-PD-1 antibody)^23^. The eligibility criteria for the patients included: (a) underwent at least one previous systemic therapy; (b) metastatic STS or bone sarcoma diagnosis histologically confirmed by pathological expert in accordance with the WHO Classification of Tumors and Soft Tissue and Bone; (c) had at least one measurable lesion by RECIST 1.1 and one biopsy accessible lesion; (d) 12 years or older; (d) at least 12 weeks of life expectancy^23^. STS group included the following subtypes: undifferentiated pleomorphic sarcoma (UPS), dedifferentiated liposarcoma (DDLPS), synovial sarcoma (SS) and leiomyosarcoma (LMS). The bone sarcoma group included: osteosarcoma (OS), Ewing sarcoma (ES) and dedifferentiated chondrosarcoma (CS). All the patients received intravenously a doss of 200 mg of pembrolizumab every three weeks. According to the protocol blood and tumor samples were to be collected before pembrolizumab treatment (receiving the name of “baseline” samples) and 8 weeks after start of treatment (receiving the name of “week-8” samples). All the research and ethical approvals and permits together with the written informed consents from all the patients were obtained prior to sample collection. In summary the 31-sarcoma patient cohort consisted of 13 patients with STS, including four synovial sarcomas (SS), four leiomyosarcomas (LMS), two dedifferentiated liposarcomas (DDLPS), three undifferentiated pleomorphic sarcomas (UPS); and 18 patients with bone sarcomas, including six Ewing sarcomas (ES), nine osteosarcomas (OS) and three dedifferentiated chondrosarcomas.

The import and analysis of the available samples, 31 baseline and seven week-8 (Supplementary Table 3), to Norway was approved by the Committee for Medical Ethics in Southeastern Norway #17866. All available samples from participating centers were collected and DNA and/or RNA purified by the trial organization and shipped to Oslo University Hospital for sequencing analysis. The data were stored and analyzed by NEC OncoImmunity in the computing infrastructure specially designed for high-level protection of sensitive personal data at the University of Oslo^77^.

### Whole exome sequencing (WES) on sarcoma tumor tissue and matched PBMCs and whole transcriptome sequencing (RNA-seq) on sarcoma tumor tissue

Whole exome libraries were prepared at the Oslo University Hospital Genomics Core Facility from 100 ng of genomic DNA using the Twist Core Exome enrichment system (Twist Bioscience) following the manufacturer’s protocol. RNA sequencing libraries were constructed using the KAPA RNA Hyper kit to generate a total RNA library, which was further captured using the Twist Core Exome probe set. Variable input amounts of RNA were used depending on the availability of material (from 20-100 ng). Exomes and RNA libraries were sequenced paired-end 2×75bp using the Illumina HiSeq 4000 instrument, and FASTQ files were generated using Illumina’s bcl2fastq conversion software.

### Screening of mutational landscape of sarcoma through a comprehensive variant calling approach

The screening and characterization of single nucleotide variants (SNVs) and small insertions and deletion (indels) sculpturing the tumor genome was conducted using NeoMutate^47^, a tool previously published by our group yielding very high validation rates^47^. After an initial thorough inspection of the raw sequencing data, including quality control and adapter trimming, the high quality paired-end reads were mapped to the human genome (GRCh38) using BWA-MEM^78^(v0.7.17-r1188). The output BAM files were treated according to the genome analysis toolkit (GATK, v4.0.6.0) best practices^79^ (including PCR duplicate marking and realignment optimization). NeoMutate incorporates an ensemble of six independent state-of-the-art somatic variant calling tools enabling the capture of the full mutational profile from tumor-normal matched WES data. Only those variant candidates detected with high confidence at least by two out of the six tools were kept in order to avoid false positive mutation calls. The variants were additionally filtered according to different quality thresholds (including minimum read depth (DP) of 10 reads for both tumor and normal data, more than three reads supporting the mutation in the tumor sample at the variant position). Ensembl Variant Effect Predictor (VEP)^80^ toolkit was used to annotate the functional effect of the detected variants on the resulting gene product. VEP was also exploited to identify the non-synonymous variants, in other words, those mutations altering the amino acid sequence of the tumor proteome, underpinning the neoantigen landscape of tumors (see Methods section “Characterization of sarcomas immunogenicity through neoantigen prediction”). Importantly, the expression of the somatic variations was evaluated using RNA-seq data, and only the expressed somatic variants were retained for the neoantigen prediction step. This is because the altered peptides need to be expressed by the tumor to result in the production of a neoantigen.

In addition to somatic variant identification, GATK-HaplotypeCaller (v4.0.6)^81^ was used for germline variant identification, and VEP^80^ for the variant annotation. Importantly, the combined effect of proximal (nearby) variants (either germline or somatic) altering the same protein, and therefore, the same neoantigenic peptide, was evaluated. Haplotype phasing is the bioinformatics process of statistical estimation of haplotypes from genotype data. WhatsHap^82^ (v0.17) was used giving as input both tumor WES and RNA-seq data to assess the phase relationship between proximal variants, in other words, to evaluate whether to nearby variants were affecting the same allele in the same tumor subclone. After the phasing, Haplosaurus^83^ (included in VEP package), was called to assess the joined functional impact of the phased variants and fully reconstruct the mutated protein sequence. Phasing step ensures that the selected neoantigen repertoire arisen from the mutated proteins correctly represent the patient’s genome, increasing the chances of anti-tumor response to immunotherapy.

### Characterization of sarcoma gene fusions

It is widely recognized that large chromosomal rearrangements and fusion genes play a critical role in underpinning and driving the sarcomagenesis course in specific morphological subtypes, making them a valuable diagnostic marker^15, 84^. The accurate identification of sarcoma fusion genes helps to understand its pathogenesis and the development of specific treatment strategies against the targetable fusions. In addition, gene fusions represent an incredibly valuable source of potentially immunogenic neoantigens that can mediate the anticancer immune response to ICI, even in those tumors with low TMB^85^. We predicted the gene fusions from RNA-seq data using Arriba^86^, the winner method for gene fusion detection in the ICGC-TCGA DREAM Somatic Mutation Calling–RNA Challenge. Arriba^86^, developed for clinical research setting, it is based in the ultrafast STAR aligner, and it computes a confidence score (low, medium or high) reflecting the likelihood of the fusion being generated due to an underlying genomic rearrangement specific to the tumor and not due to a sequencing artefact. Low confidence gene fusions were discarded from downstream analysis.

### Tumor mutation burden (TMB) calculation

TMB was defined as the number of non-synonymous somatic SNV and indels with a VAF of at least %5 per megabase in the coding area of the cancer genome, as recommended by the guidelines of the Friends of Cancer Research TMB Harmonization Project^87^. Fusions, CNVs, non-coding and synonymous mutations were discarded for TMB calculation.

### Human Leukocyte Antigen (HLA) typing

HLA alleles of each patient were inferred in silico using OncoHLA^88^ providing peripheral blood mononuclear cell (PBMC) WES data as input. OncoHLA uses an integer linear programming algorithm together with prior probabilities of the allelic ethnic frequencies to determine the closest-matched HLA allele from the IPD-IMGT/HLA Database^89^ (v.3.41.2). The output includes the HLA types for both class I and class II up to four field of resolution and the associated HLA gene, transcript and protein sequences.

### HLA expression quantification and HLA somatic variant screening in sarcoma samples

It is well established that cancer cells can exploit several HLA-associated immune evasion mechanisms to hijack immune system^90, 91^. A comprehensive scrutiny of the HLA status in the tumor was conducted using a previously developed method by our group^92^. Using the typed HLA alleles, an exhaustive profiling of the somatic mutations affecting each individual HLA allele was carried, and their functional impact in the corresponding HLA protein sequences was annotated. In addition, the expression (abundance), reported as transcripts per million mapped reads (TPM), of each allele was quantified by mapping the RNA-seq reads to the inferred HLA sequences, to evaluate whether any allele was downregulated – a well-known immune escape mechanism in tumor development.

### Isoform and gene-level expression quantification using RNA-seq data from sarcoma samples

Gene isoform expression profiling from the RNA-seq data of both STS and bone sarcoma was carried utilizing Kallisto^93^ (v0.43.1), based on pseudoalignments of the reads and expectation-maximization (EM) algorithm to conduct isoform-level expression quantification. The reference transcriptome for GRCh38 genome, required as input, was obtained from Ensembl database version 95. Kallisto reports the abundance transcript level measured TPM.

The expression values of each transcript were used for several analyses that will be further detailed in the following sections, including: (1) Calculation of the abundance of the potential neoantigens generated from the proteins affected by non-synonymous variants in the tumor; (2) Differential expression and enrichment analysis; (3) TME profiling; (4) Manual inspection of immune-related genes expression.

### Characterization of sarcomas immunogenicity through neoantigen prediction analysis

The immunogenicity of the mutated peptides derived from tumor-specific alterations was assessed using NEC Immune Profiler (NIP) modular neoantigen pipeline developed by NEC OncoImmunity, comprising several proprietary T cell epitope machine-learning (ML) prediction algorithms. Neoantigen predictions for HLA-A and -B were conducted for each patient with peptides of length 9 and 10. Due to the lack of HLA-C validated data influencing the accuracy of the ML models, HLA-C was not evaluated. The pipeline considers several features determining the immunogenicity of a neoantigen, including:

1. The binding affinity of the peptide for the MHC/HLA molecule. NIP exploits three distinct binding affinity ML predictors that compute IC50 (nM) scores for each mutated peptide. The lower the IC50 score, the stronger the binding of the peptide to the specific HLA molecule.
2. The peptide’s ability to be efficiently digested by the antigen processing machinery (APM). An ensemble of 13 of Support Vector Machines (SVM) included in NIP and trained on validated mass spectrometry immunopeptidome datasets determine which peptides have the optimal features to be efficiently processed by the APM, which include cleavage probability by the proteasome and antigen processing transport (TAP) efficiency.
3. The expression level of the mutated peptide. The expression of each neoantigen candidate was computed by summing the expression values (TPMs) of all the isoforms coding for the specific peptides under consideration, which is critical for accurate prediction of immunogenic neoantigens. To determine the specific abundance of the mutated peptide, the sum of the expression levels of all the isoforms containing the peptide was adjusted according to the variant allele frequency (VAF) computed at RNA level.
4. The relative uniqueness of the candidate neoantigens compared to the normal peptides of healthy tissue was evaluated to avoid cross-reaction with self-antigen sequences.

The final result is summarized in a single score ranging from 0 to 1, known as the antigen presentation score (AP), which indicates the T cell recognition probability, ranging from 0 to 1, with 1 signifying the highest likelihood that the given neoantigen is immunogenic. Neoantigen load (NAL) was computed by calculating the number of neoantigens with an AP higher than 0.5. Neoantigen quality in this study was defined as the mean AP score for the top ten neoantigen candidates ranked by AP score.

### Tumor microenvironment (TME) profiling

QuanTIseq^94^ deconvolution algorithm was utilized to analyze the immune cell composition of each sarcoma sample. QuanTIseq has demonstrated a robust overall performance^95^ and is one of the very few methods generating an absolute score can be interpreted as a cell fraction, which allows both inter- and intra-sample comparisons. It takes as input RNA-seq reads and quantifies via deconvolution of cell fractions based on constrained least squares regression the proportion of ten different immune cell phenotypes, including B cells, M1 and M2 macrophages, CD8+, CD4+ and regulatory T cells, natural killer cells (NK), among others.

In addition to QuanTIseq, TcellExTRECT^96^ R package was applied to estimate infiltrated T cell fraction. The tool employs WES data and makes use of a signal based on somatic copy number from V(D)J recombination to directly quantify the proportion of T cells.

Further, the ESTIMATE algorithm was applied to calculate the stromal and immune scores for each sarcoma sample using the normalized gene expression values as input^97^.

### Selection of immune-related genes

We conducted an exhaustive literature research and compiled a list of 282 genes known to be related with immune system interaction and response^98–100^. The full immune-related gene list is provided in Supplementary Table 4.

### Differential expression (DE) analysis and enrichment analysis

In order to characterize the differentially expressed genes (DEG) between week-8 and baseline samples, the DESeq2^101^ (v1.30.1) R package was selected. “Tximport”^102^ (1.18.0) R package was utilized to import the transcript-level expression estimates generated by Kallisto and produce gene-level count matrices and normalizing offsets, as required by DeSeq2^101^. DeSeq2^101^ models the counts using normalization factors to account for differences in the library depth, estimates the gene-wise dispersions and uses shrinkage of effect size to remove the low count genes. Then, it fits a negative binomial model and performs Wald test hypothesis testing. DEGs were obtained by applying a filter of p-value<0.05.

The results of the DE analysis were expanded by conducting an enrichment analysis using an overrepresentation analysis (ORA) to associate the expression data with specific biological processes (BP). The R package called “goseq”^103^ (1.42.0) conducted ORA of Gene Ontology (GO) terms, which corrects the results based on gene length bias of DEGs. DEGs were first separated into up and down-regulated genes attending to their fold change (FC) (greater than 1 or smaller than −1, for up and down-regulated genes, respectively), and then goseq^103^ was used to detect overrepresented up and down-regulated BPs.

### Statistical analysis

The statistical analyses in this study were conducted using R (4.0.3) and python (3.8) programming languages. The criteria for the annotation of the different determinants of each patient to ICI therapy, such as progression-free survival (PFS) and overall survival (OS), has been previously described in^23^. Survival curves were plotted using the Kaplan–Meier (KM) functionality within “lifelines” python library to compare PFS for a different set of individual covariates (univariate analysis) and in conjunction (multivariate analysis). Differences in median PFS were assessed using the log-rank test and multivariate long-rank test.

### Supervised optimal binning

Kaplan-Meier (KM) survival analysis entails splitting individuals into two or more groups and comparing survival times between individuals in the groups. For a numeric covariate, a threshold is required to perform such a split; that is, we must select a threshold to bin individuals into groups. Prior work^104^ has shown that for such binning problems, the only thresholds which change group composition are the values which actually occur in the dataset. For example, when splitting individuals into groups based on TMB, the only TMB thresholds which change to which group individuals are assigned are the actual TMB values of the individuals. Thus, we determined an “optimal” threshold with respect to KM analysis by evaluating the log rank test p-value for all possible binnings of individuals in the dataset; we implemented this by simply trying each observed value for the numeric covariate of interest in the patient dataset. Only threshold cutoffs generating bins with at least 10 individuals each were evaluated.

### Data availability

The NGS data are stored at the “Services for sensitive data” of the University of Oslo Computing Centre, providing strong protection but also controlled access for approved users in compliance with the patient consents. Access may be obtained by contacting OM or LAMZ.

## Acknowledgements

This work was supported by grant MISP # 53061 from Merck and from the Norwegian Research Council. We are grateful for the services of the OUH Genomics Core Facility (oslo.genomics.no)^a^ and the UiO Services for sensitive data^b^, and for NN and MM in collecting, preparing and shipping DNA and RNA samples.

## Footnotes

a https://genomics.no

b https://www.uio.no/english/services/it/research/sensitive-data/about/index.html

